# Characterising post-COVID syndrome more than 6 months after acute infection in adults; prospective longitudinal cohort study, England

**DOI:** 10.1101/2021.03.18.21253633

**Authors:** Zahin Amin-Chowdhury, Ross J Harris, Felicity Aiano, Maria Zavala, Marta Bertran, Ray Borrow, Ezra Linley, Shazaad Ahmad, Ben Parker, Alex Horsley, Bassam Hallis, Jessica Flood, Kevin E Brown, Gayatri Amirthalingam, Mary E Ramsay, Nick Andrews, Shamez N Ladhani

**Affiliations:** Immunisation and Countermeasures Division, Public Health England, Colindale, London NW9 5EQ, UK; Statistics, Modelling, and Economics Department, Public Health England, Colindale, London NW9 5EQ, UK; Sero-epidemiology Unit, Public Health England, Public Health Laboratory Manchester, Manchester Medical Microbiology Partnership, Manchester Royal Infirmary, Oxford Road, Manchester, M13 9WL; Department of Virology, Manchester Medical Microbiology Partnership, Manchester Foundation Trust, Manchester Academic Health Sciences Centre, Manchester, UK; The NIHR Manchester Clinical Research Facility, Manchester University NHS Foundation Trust, Manchester, UK; Kellgren Centre for Rheumatology, NIHR Manchester Biomedical Research Centre, Manchester University NHS Foundation Trust, Manchester, UK; Division of Infection, Immunity and Respiratory Medicine, University of Manchester, Manchester, UK; Immunoassay Lab, National Infection Service, Porton Down SP4 0JG; Virus Reference Department, Reference Microbiology, National Infection Service, Public Health England Colindale. 61 Colindale Avenue, London NW9 5EQ; Paediatric Infectious Diseases Research Group (PIDRG), St. George’s University of London, Cranmer Terrace, London SW17 0RE, UK

**Keywords:** long COVID, prospective cohort, control group

## Abstract

**Background:** Most individuals with COVID-19 will recover without sequelae, but some will develop long-term multi-system impairments. The definition, duration, prevalence and symptoms associated with long COVID, however, have not been established.

**Methods:** Public Health England (PHE) initiated longitudinal surveillance of clinical and non-clinical healthcare workers for monthly blood sampling for SARS-CoV-2 antibodies in March 2020. Eight months after enrolment, participants completed an online questionnaire including 72 symptoms in the preceding month. Symptomatic mild-to-moderate cases with confirmed COVID-19 were compared with asymptomatic, seronegative controls. Multivariable logistic regression was used to identify independent symptoms associated with long COVID.

**Results:** All 2,147 participants were contacted and 1,671 (77.8%) completed the questionnaire, including 140 (8.4%) cases and 1,160 controls. At a median of 7.5 (IQR 7.1-7.8) months after infection, 20 cases (14.3%) had ongoing (4/140, 2.9%) or episodic (16/140, 11.4%) symptoms. We identified three clusters of symptoms associated with long COVID, those affecting the sensory (ageusia, anosmia, loss of appetite and blurred vision), neurological (forgetfulness, short-term memory loss and confusion/brain fog) and cardiorespiratory (chest tightness/pain, unusual fatigue, breathlessness after minimal exertion/at rest, palpitations) systems. The sensory cluster had the highest association with being a case (aOR 5.25, 95% CI 3.45-8.01). Dermatological, gynaecological, gastrointestinal or mental health symptoms were not significantly different between cases and controls.

**Conclusions:** Most persistent symptoms reported following mild COVID-19 were equally common in cases and controls. While all three clusters identified had a strong association with previous COVID-19 infection, the sensory cluster had the highest specificity and strength of association.

**Key points:** Compared to controls, we identified three clusters of symptoms affecting the sensory, neurological and cardiorespiratory systems that were more prevalent among cases. Notably, gastrointestinal and dermatological symptoms and symptoms related to mental health were as prevalent among cases as controls.

## Introduction

The clinical manifestations of severe acute respiratory syndrome coronavirus 2 (SARS-CoV- 2) – the virus responsible for coronavirus disease 19 (COVID-19) – range from asymptomatic infection to severe disease leading to multi-organ failure and death.^1, 2^ While for many survivors, the infection is mild and short-lived, with emerging evidence, it is now recognised that an as-yet undefined proportion will go on to develop persistent symptoms after recovering from their infection.^3^ Various terms have been used to describe the condition, including ‘post-acute COVID-19’, ‘post-COVID-19 syndrome’ and ‘long COVID’.^4, 5^ In the UK, NICE–SIGN–RCGP have developed guidelines to support patients with long COVID, but the definition for this condition is very non-specific, and includes any individual with signs and symptoms that develop during or after an infection consistent with COVID-19, continuing for more than 12 weeks and are not explained by an alternative diagnosis.^6^

The reported symptoms in individuals recovering from COVID-19 are varied and may manifest as episodic, cyclical or persistent. Symptoms affecting every organ system have been described, particularly affecting the cardiovascular, respiratory and neuromuscular system, as well as non-specific symptoms such as excessive fatigue, body aches and “brain fog”.^7, 8^ Chronic fatigue has also been widely reported in some patients recovering from COVID-19, which is not surprising given that this syndrome can occur after any infection.^7, 8^ Currently, our understanding of long COVID-19 comes from information provided by patients with COVID-19, usually those who were hospitalised and, therefore, experienced more severe disease.^9, 10^ Additionally, published studies have been limited self-selected cohorts of patients and lack of appropriate control groups to objectively assess the relative frequency of reported symptoms, which have led to surprisingly high estimates of long COVID prevalence estimates, with a pooled estimate of 80% in one systematic review. Some of the >170 symptoms reportedly caused by COVID-19 have low biological plausibility but cannot be excluded without knowledge of their frequency in the general population.^5, 11^

At the start of the pandemic, Public Health England (PHE) initiated active prospective surveillance of more than 2,000 clinical and non-clinical healthcare workers to collect monthly blood samples for SARS-CoV-2 antibody testing.^12^ Around 12% developed COVID- 19, mainly during March/April 2020. In order to better understand the characteristics of long COVID in healthy adults with mild-to moderate COVID-19, we asked all participants to complete an online questionnaire about their physical and mental state in November 2020. Knowledge of their SARS-CoV-2 RT-PCR and antibody status allowed us to accurately identify laboratory-confirmed symptomatic COVID-19 cases along with a large control group of asymptomatic adults who remained SARS-CoV-2 antibody negative throughout the surveillance period. Our objective was to estimate the prevalence of persistent symptoms in working-age adults with confirmed COVID-19. We also compared reported long COVID symptoms identified through a literature review and mental health state of individuals with confirmed COVID-19 more than 6 months after their infection with the asymptomatic, SARS-CoV-2 antibody negative controls.

## Methods

### Study design and participants

In March 2020, PHE initiated a prospective cohort study (ESCAPE) of PHE and National Health Service (NHS) healthcare workers to collect monthly blood samples for SARS-CoV-2 antibodies at multiple sites in England (PHE London, PHE Porton Down, PHE Manchester, NHS Manchester Royal Infirmary and NHS Wythenshawe Hospital).^12^ Participants included office-based workers, laboratory workers, as well as clinical patient-facing and non-patient facing healthcare workers, most of whom working on-site throughout national lockdown.Participants were provided with information about the study and provided written informed consent prior to enrolment. A short online questionnaire requesting information about any illness, symptoms and SARS-CoV-2 testing since the previous blood sampling visit was completed on-site by the participant at each blood sampling visit. If participants missed a visit, they could still continue to take part at the next visit.^13^

### Questionnaire design

To assess any long-term effects of COVID-19, participants were invited to complete a detailed online questionnaire in November 2020. The questionnaire was developed based on all possible symptoms associated with long COVID that were reported online or in the scientific literature.^14, 15^ The questionnaire was piloted among adults with and without COVID-19 and revised according to feedback received. The final questionnaire (**Supplement 1**) included questions on demographics, COVID-19 symptoms, PCR testing, chronic health conditions as well as whether they or not they had experienced any of 72 symptoms in the month prior to questionnaire completion. Symptoms were sub-grouped into (1) neurological, (2) dermatological, (3) sensory, (4) respiratory, (5) gastrointestinal, (6) cardiovascular, (7) mental health, and (8) other. The questionnaire was disseminated to ESCAPE participants electronically using the Snap Professional 11 platform on 03 November 2020. Participants received regular email reminders to complete the questionnaire until the survey closed on 12 February 2021.

### Laboratory methods

The laboratory methods for SARS-CoV-2 antibody testing are described elsewhere.^12^ In brief, blood samples were tested using five different assays for Nucleoprotein (Roche N and Abbott) and for Spike (EUROIMMUN, Roche S, and an in-house assay named RBD) antibodies. A participant was seropositive if positive for Nucleoprotein or Spike antibodies in any assay. Indeterminate results and insufficient samples were excluded.

### Definitions

A case was defined as any participant with COVID-19 symptoms and laboratory-confirmed positive SARS-CoV-2 antibodies with an interval of at least 6 months from their first positive SARS-CoV-2 RT-PCR test or, if not tested, the onset of their COVID-19 symptoms until questionnaire completion. The uninfected control group comprised SARS-CoV-2 antibody negative participants who remained asymptomatic until questionnaire completion. Participants who became SARS-CoV-2 RT-PCR or antibody positive within 6 months of questionnaire completion were not included in the analysis, as were those with COVID-19 symptoms who were SARS-CoV-2 antibody negative >30 days after their illness and those who developed COVID-19 symptoms after their last negative antibody test.

### Data management and statistical analyses

Data were managed using Microsoft Access (Microsoft Corporation, Redmond, Washington) and analysed using Stata 15.1 (StataCorp LLC, College Station, Texas). Participant characteristics were described as counts and proportions. Non-normally distributed continuous variables are presented as medians with interquartile range. Categorical data are presented as a frequency (percentage) and compared using χ^2^, Fisher’s exact or Kruskal-Wallis tests as appropriate, unless otherwise stated.

Logistic regression models were fitted for symptoms identified as statistically significant (p<0.05) in the univariable χ^2^, or Fisher’s exact test, as appropriate and adjusted for age group, sex, comorbidity status, ethnicity and workplace role. Because of multiple comparisons, a p value of ≤0.001 was considered statistically significant. Symptoms with less than 5% reported in cases and controls were also excluded due to being extremely rare. Potential interactions between covariates were assessed using likelihood ratio tests. The controls were compared with antibody positive participants which were excluded to assess for significant difference. The results are reported as odd ratios (OR) and adjusted odds ratios (aOR) with 95% confidence intervals (CI). Symptoms within organ systems of both clinical and statistical significance were regrouped and fitted into logistic regression models adjusting for covariates described above.^16^

## Results

### Participant characteristics

All 2,147 ESCAPE participants were contacted by email and 1,671 (77.8%) completed the online questionnaire. Asymptomatic, antibody positive participants (n=73), those who developed COVID-19 within 6 months of completing the questionnaire (n=172) and those reporting COVID-19 symptoms or a positive RT-PCR result who remained SARS-CoV-2 antibody negative more than 30 days after their illness (n=126) were excluded (**Figure 1**). There were, therefore, 140 confirmed cases (10.8%), including 37 with PCR-confirmed COVID-19 and 103 with COVID-19 symptoms who had not been tested by RT-PCR. The median interval between illness and questionnaire completion in cases was 7.5 months (IQR 7.1-7.8). The control group comprised of 1,160 (89.2%) participants who remained asymptomatic and antibody negative throughout the surveillance period.

**Figure 1.**
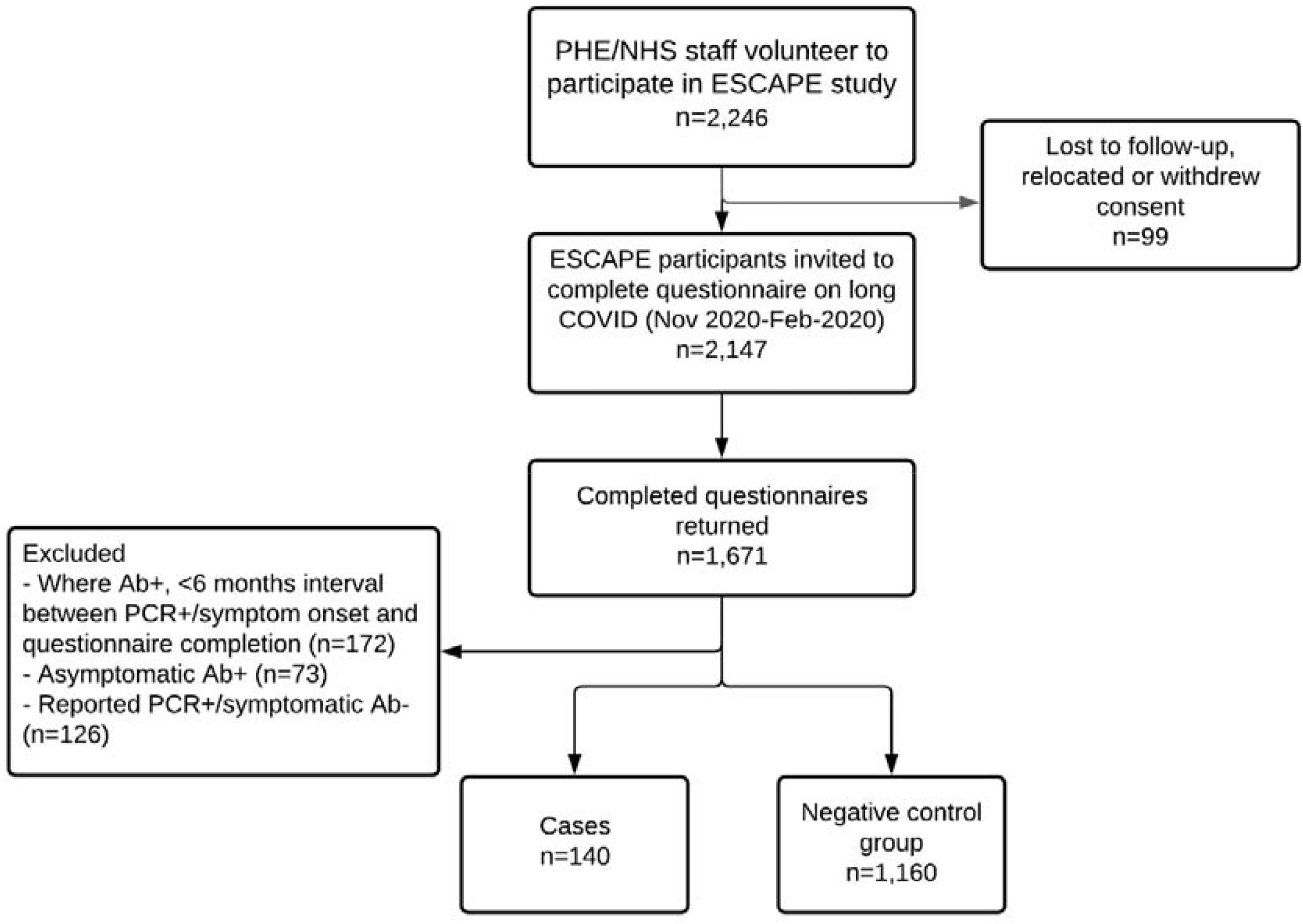
Flowchart of participation in the study and selection of cases and uninfected control group.

Participant characteristics are described in **Table 1**. The median age of cases was 41 (IQR 31-52) years, which was similar to the control group (42 [33-53]; p=0.12), as was the proportion who were female and those with underlying comorbidities. Workplace setting varied between cases and controls, with patient-facing hospital staff comprising a higher proportion of cases than controls (51.7% vs 23.0%). None of the cases were hospitalised for COVID-19.

**Table 1.** Characteristics of cohort of cases and uninfected control group.

|  | **Cases**<br>**N (%)**<br>**n=140** | **Control group**<br>**N (%)**<br>**n=1,160** | **Total**<br>**N (%)**<br>**n=1300** |
| --- | --- | --- | --- |
| **Age, median (IQR)** | 41 (31-52) | 42 (33-52) | 43 (33-53) |
| **Age group** |  |  |  |
| 20-29 | 29 (20.3%) | 177 (15.2%) | 206 (15.8%) |
| 30-39 | 34 (23.8%) | 282 (24.2%) | 316 (24.2%) |
| 40-49 | 39 (27.3%) | 314 (27.0%) | 353 (27.0%) |
| 50-59 | 35 (24.5%) | 284 (24.4%) | 319 (24.4%) |
| 60+ | 6 (4.2%) | 107 (9.2%) | 113 (8.6%) |
| **Sex** |  |  |  |
| Male | 41 (28.7%) | 362 (31.1%) | 403 (30.8%) |
| Female | 102 (71.3%) | 802 (68.9%) | 904 (69.2%) |
| **Ethnicity** |  |  |  |
| White | 108 (76.1%) | 994 (85.8%) | 1102 (84.7%) |
| Black African/Caribbean | 7 (4.9%) | 26 (2.2%) | 33 (2.5%) |
| Indian sub-continent | 12 (8.5%) | 75 (6.5%) | 87 (6.7%) |
| Mixed | 4 (2.8%) | 13 (1.1%) | 17 (1.3%) |
| Other | 11 (7.7%) | 51 (4.4%) | 62 (4.8%) |
| **Workplace setting** |  |  |  |
| Hospital with regular patient contact | 74 (51.7%) | 268 (23.0%) | 342 (26.2%) |
| Hospital with occasional patient contact | 10 (7.0%) | 114 (9.8%) | 124 (9.5%) |
| Hospital with no patient contact | 13 (9.1%) | 137 (11.8%) | 150 (11.5%) |
| Office-based/PHE laboratory | 33 (23.1%) | 453 (38.9%) | 486 (37.2%) |
| Working from home | 6 (4.2%) | 133 (11.4%) | 139 (10.6%) |
| Other | 7 (4.9%) | 59 (5.1%) | 66 (5.0%) |
| **Symptom group** |  |  |  |
| Ongoing | 5 (3.5%) | - | 5 (0.4%) |
| Episodic | 16 (11.2%) | - | 16 (1.2%) |
| Well | 122 (85.3%) | 1164 (100.0%) | 1286 (98.4%) |
| **Has one or more comorbidities** | 26 (18.4%) | 133 (11.6%) | 159 (12.4%) |
| **Hospitalised for COVID-19** | 0 (0.0%) | - | 0 (0.0%) |

### Persisting symptoms

Most participants reported that they recovered without persisting symptoms, but 14.7% (20/140) had ongoing (4/140, 2.9%) or episodic (16/140, 11.4%) symptoms after COVID-19 (**Table 1**). The four participants with ongoing symptoms were female, none had underlying comorbidities and all reported unusual fatigue and breathlessness after minimal exertion in the month prior to questionnaire completion, with three also reporting confusion, forgetfulness, muscle aches and anxiety. Of the 16 cases with episodic symptoms, 12 (75.0%) were female and 2/16 (12.5%) had at least one or more comorbidities. All 12 reported experiencing stress in the previous month, and more than three-quarters (75-94%) reported problems sleeping through the night, mood swings, forgetfulness, unusual fatigue, muscle aches or confusion/brain fog.

### Cases versus controls

In the univariable analysis, 72 symptoms were compared in cases and controls (**Table 2**). The controls did not present with symptoms significantly different to antibody positive participants which were excluded from the analyses. Neurological symptoms of statistical significance included problems sleeping through the night (60.7% vs 51.5%), forgetfulness (35.0% vs 19.0%), confusion/brain fog/trouble focussing attention (20.7% vs 14.7%), trouble trying to form words (15.7% vs 9.2%), short-term memory loss (20.7% vs 5.6%) and, less frequently, difficulty swallowing (6.4% vs 2.4%), twitching of fingers and toes (5.7% vs 2.4%) and trembling (5.7% vs 1.7%). Respiratory symptoms of interest included unusual fatigue/tiredness after exertion (39.3% vs 17.5%), breathlessness after minimal exertion (25.7% vs 10.2%), chest tightness/pain (18.6% vs 8.2%), fits of coughing (13.6% vs 6.5%) and breathlessness at rest (9.3% vs 2.8%). Sensory symptoms included anosmia (18.6% vs 0.8%), ageusia (17.1% vs 0.6%), loss of appetite (14.3% vs 5.8%) and blurred vision (13.6% vs 6.9%). Palpitations was the only cardiovascular symptom of statistical significance (21.4% vs 11.5%). There were no significant symptoms within dermatological, gynaecological, gastrointestinal or mental health sub-groups. Rare symptoms (prevalence <5% in cases) of statistical significance including slurred speech, hallucinations and collapse were excluded from further analyses (**Table 2**).

**Table 2.**
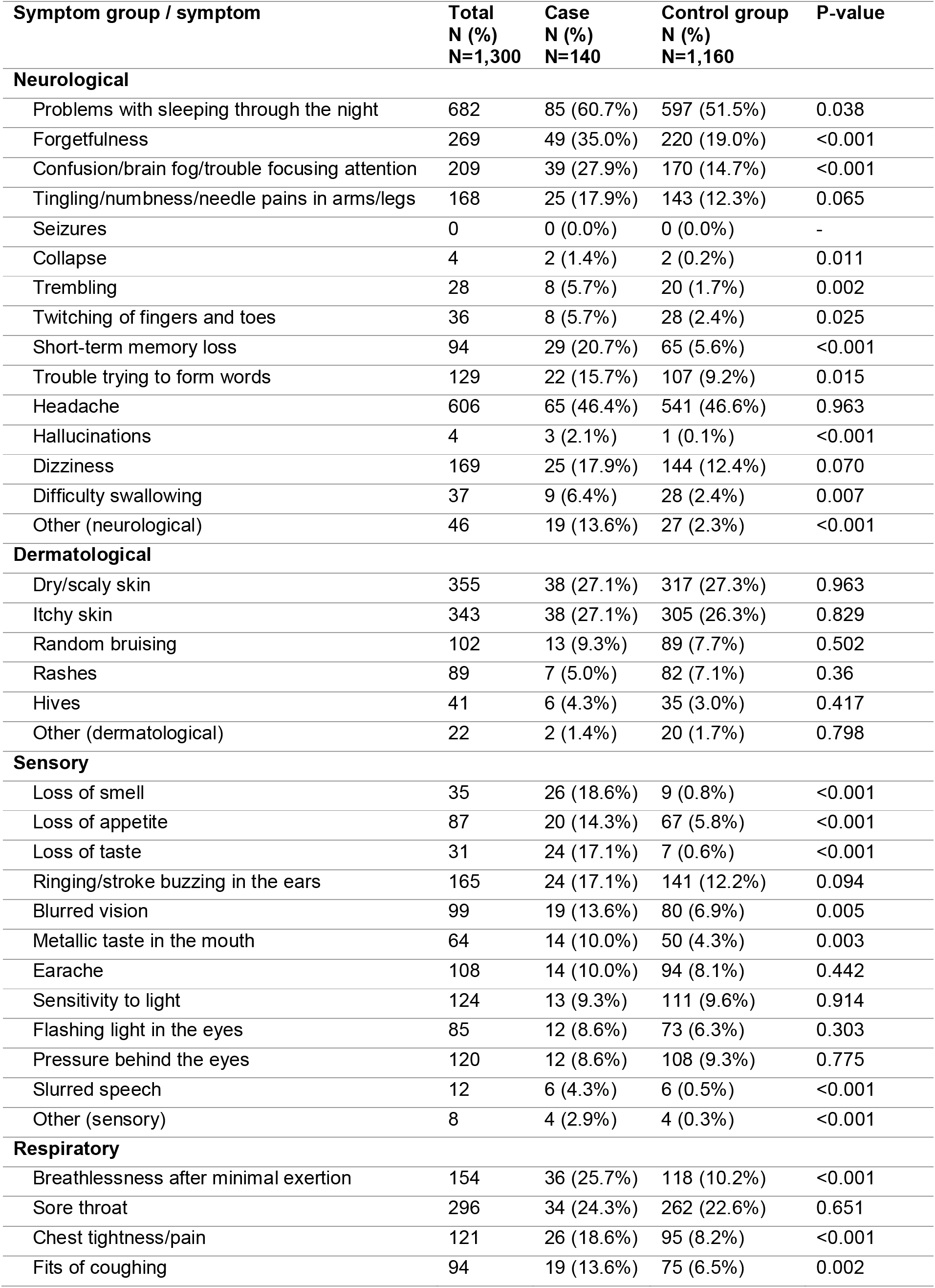
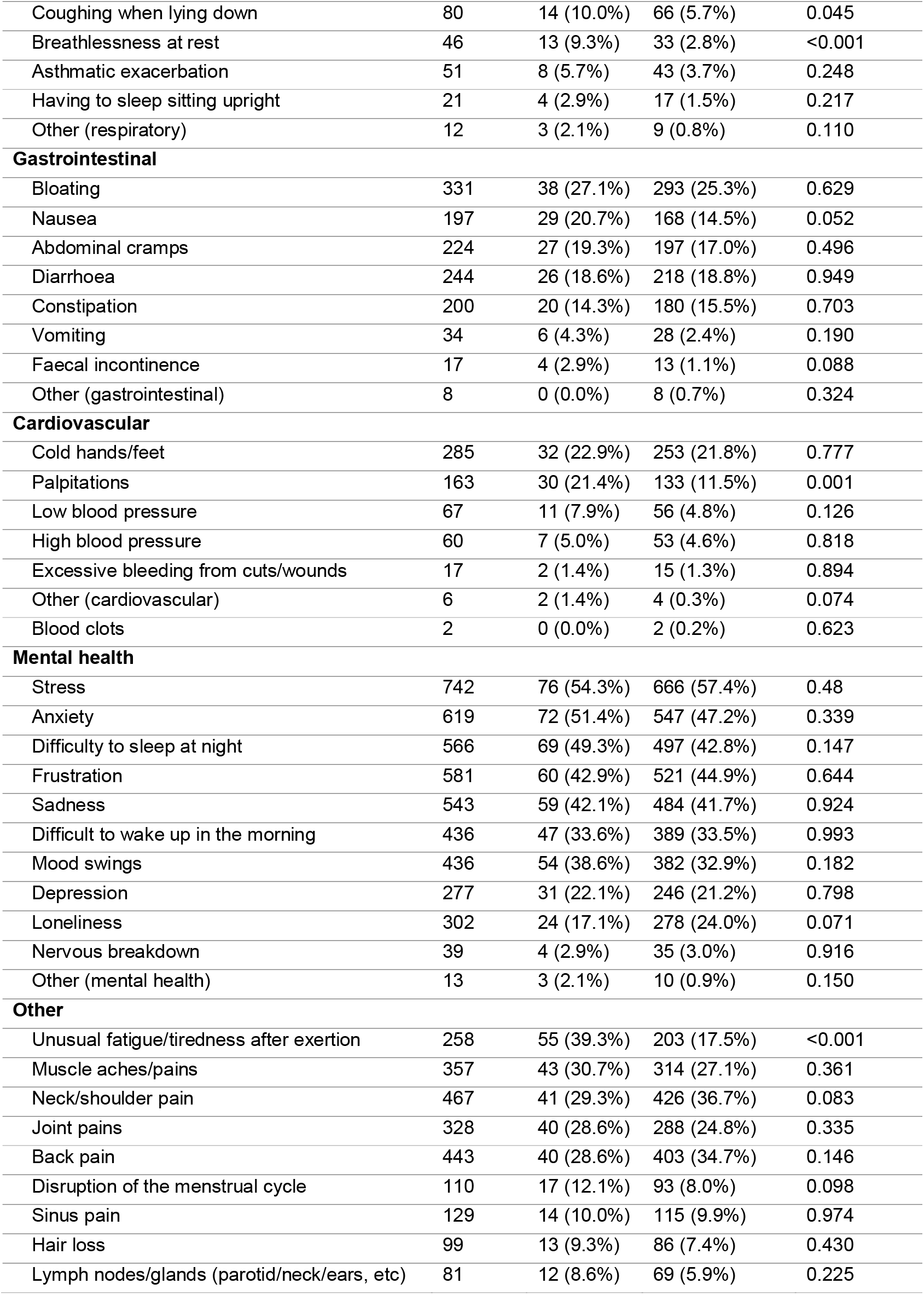
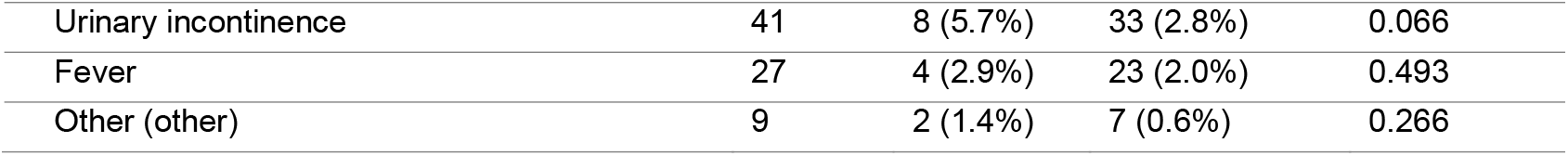
Univariable analysis of symptoms by infection status (case, control group)

| Symptom group / symptom | Total<br>N (%)<br>N=1,300 | Case<br>N (%)<br>N=140 | Control group<br>N (%)<br>N=1,160 | P-value |
| --- | --- | --- | --- | --- |
| **Neurological** |  |  |  |  |
| Problems with sleeping through the night | 682 | 85 (60.7%) | 597 (51.5%) | 0.038 |
| Forgetfulness | 269 | 49 (35.0%) | 220 (19.0%) | <0.001 |
| Confusion/brain fog/trouble focusing attention | 209 | 39 (27.9%) | 170 (14.7%) | <0.001 |
| Tingling/numbness/needle pains in arms/legs | 168 | 25 (17.9%) | 143 (12.3%) | 0.065 |
| Seizures | 0 | 0 (0.0%) | 0 (0.0%) | - |
| Collapse | 4 | 2 (1.4%) | 2 (0.2%) | 0.011 |
| Trembling | 28 | 8 (5.7%) | 20 (1.7%) | 0.002 |
| Twitching of fingers and toes | 36 | 8 (5.7%) | 28 (2.4%) | 0.025 |
| Short-term memory loss | 94 | 29 (20.7%) | 65 (5.6%) | <0.001 |
| Trouble trying to form words | 129 | 22 (15.7%) | 107 (9.2%) | 0.015 |
| Headache | 606 | 65 (46.4%) | 541 (46.6%) | 0.963 |
| Hallucinations | 4 | 3 (2.1%) | 1 (0.1%) | <0.001 |
| Dizziness | 169 | 25 (17.9%) | 144 (12.4%) | 0.070 |
| Difficulty swallowing | 37 | 9 (6.4%) | 28 (2.4%) | 0.007 |
| Other (neurological) | 46 | 19 (13.6%) | 27 (2.3%) | <0.001 |
| **Dermatological** |  |  |  |  |
| Dry/scaly skin | 355 | 38 (27.1%) | 317 (27.3%) | 0.963 |
| Itchy skin | 343 | 38 (27.1%) | 305 (26.3%) | 0.829 |
| Random bruising | 102 | 13 (9.3%) | 89 (7.7%) | 0.502 |
| Rashes | 89 | 7 (5.0%) | 82 (7.1%) | 0.36 |
| Hives | 41 | 6 (4.3%) | 35 (3.0%) | 0.417 |
| Other (dermatological) | 22 | 2 (1.4%) | 20 (1.7%) | 0.798 |
| **Sensory** |  |  |  |  |
| Loss of smell | 35 | 26 (18.6%) | 9 (0.8%) | <0.001 |
| Loss of appetite | 87 | 20 (14.3%) | 67 (5.8%) | <0.001 |
| Loss of taste | 31 | 24 (17.1%) | 7 (0.6%) | <0.001 |
| ringing/stroke buzzing in the ears | 165 | 24 (17.1%) | 141 (12.2%) | 0.094 |
| Blurred vision | 99 | 19 (13.6%) | 80 (6.9%) | 0.005 |
| Metallic taste in the mouth | 64 | 14 (10.0%) | 50 (4.3%) | 0.003 |
| Earache | 108 | 14 (10.0%) | 94 (8.1%) | 0.442 |
| Sensitivity to light | 124 | 13 (9.3%) | 111 (9.6%) | 0.914 |
| Flashing light in the eyes | 85 | 12 (8.6%) | 73 (6.3%) | 0.303 |
| Pressure behind the eyes | 120 | 12 (8.6%) | 108 (9.3%) | 0.775 |
| Slurred speech | 12 | 6 (4.3%) | 6 (0.5%) | <0.001 |
| Other (sensory) | 8 | 4 (2.9%) | 4 (0.3%) | <0.001 |
| **Respiratory** |  |  |  |  |
| Breathlessness after minimal exertion | 154 | 36 (25.7%) | 118 (10.2%) | <0.001 |
| Sore throat | 296 | 34 (24.3%) | 262 (22.6%) | 0.651 |
| Chest tightness/pain | 121 | 26 (18.6%) | 95 (8.2%) | <0.001 |
| Fits of coughing | 94 | 19 (13.6%) | 75 (6.5%) | 0.002 |
| Coughing when lying down | 80 | 14 (10.0%) | 66 (5.7%) | 0.045 |
| Breathlessness at rest | 46 | 13 (9.3%) | 33 (2.8%) | <0.001 |
| Asthmatic exacerbation | 51 | 8 (5.7%) | 43 (3.7%) | 0.248 |
| Having to sleep sitting upright | 21 | 4 (2.9%) | 17 (1.5%) | 0.217 |
| Other (respiratory) | 12 | 3 (2.1%) | 9 (0.8%) | 0.110 |
| **Gastrointestinal** |  |  |  |  |
| Bloating | 331 | 38 (27.1%) | 293 (25.3%) | 0.629 |
| Nausea | 197 | 29 (20.7%) | 168 (14.5%) | 0.052 |
| Abdominal cramps | 224 | 27 (19.3%) | 197 (17.0%) | 0.496 |
| Diarrhoea | 244 | 26 (18.6%) | 218 (18.8%) | 0.949 |
| Constipation | 200 | 20 (14.3%) | 180 (15.5%) | 0.703 |
| Vomiting | 34 | 6 (4.3%) | 28 (2.4%) | 0.190 |
| Faecal incontinence | 17 | 4 (2.9%) | 13 (1.1%) | 0.088 |
| Other (gastrointestinal) | 8 | 0 (0.0%) | 8 (0.7%) | 0.324 |
| **Cardiovascular** |  |  |  |  |
| Cold hands/feet | 285 | 32 (22.9%) | 253 (21.8%) | 0.777 |
| Palpitations | 163 | 30 (21.4%) | 133 (11.5%) | 0.001 |
| Low blood pressure | 67 | 11 (7.9%) | 56 (4.8%) | 0.126 |
| High blood pressure | 60 | 7 (5.0%) | 53 (4.6%) | 0.818 |
| Excessive bleeding from cuts/wounds | 17 | 2 (1.4%) | 15 (1.3%) | 0.894 |
| Other (cardiovascular) | 6 | 2 (1.4%) | 4 (0.3%) | 0.074 |
| Blood clots | 2 | 0 (0.0%) | 2 (0.2%) | 0.623 |
| **Mental health** |  |  |  |  |
| Stress | 742 | 76 (54.3%) | 666 (57.4%) | 0.48 |
| Anxiety | 619 | 72 (51.4%) | 547 (47.2%) | 0.339 |
| Difficulty to sleep at night | 566 | 69 (49.3%) | 497 (42.8%) | 0.147 |
| Frustration | 581 | 60 (42.9%) | 521 (44.9%) | 0.644 |
| Sadness | 543 | 59 (42.1%) | 484 (41.7%) | 0.924 |
| Difficult to wake up in the morning | 436 | 47 (33.6%) | 389 (33.5%) | 0.993 |
| Mood swings | 436 | 54 (38.6%) | 382 (32.9%) | 0.182 |
| Depression | 277 | 31 (22.1%) | 246 (21.2%) | 0.798 |
| Loneliness | 302 | 24 (17.1%) | 278 (24.0%) | 0.071 |
| Nervous breakdown | 39 | 4 (2.9%) | 35 (3.0%) | 0.916 |
| Other (mental health) | 13 | 3 (2.1%) | 10 (0.9%) | 0.150 |
| **Other** |  |  |  |  |
| Unusual fatigue/tiredness after exertion | 258 | 55 (39.3%) | 203 (17.5%) | <0.001 |
| Muscle aches/pains | 357 | 43 (30.7%) | 314 (27.1%) | 0.361 |
| Neck/shoulder pain | 467 | 41 (29.3%) | 426 (36.7%) | 0.083 |
| Joint pains | 328 | 40 (28.6%) | 288 (24.8%) | 0.335 |
| Back pain | 443 | 40 (28.6%) | 403 (34.7%) | 0.146 |
| Disruption of the menstrual cycle | 110 | 17 (12.1%) | 93 (8.0%) | 0.098 |
| Sinus pain | 129 | 14 (10.0%) | 115 (9.9%) | 0.974 |
| Hair loss | 99 | 13 (9.3%) | 86 (7.4%) | 0.430 |
| Lymph nodes/glands (parotid/neck/ears, etc) | 81 | 12 (8.6%) | 69 (5.9%) | 0.225 |
| Urinary incontinence | 41 | 8 (5.7%) | 33 (2.8%) | 0.066 |
| Fever | 27 | 4 (2.9%) | 23 (2.0%) | 0.493 |
| Other (other) | 9 | 2 (1.4%) | 7 (0.6%) | 0.266 |

Other noteworthy associations, in the logistic regression model, include that unusual fatigue and problems sleeping through the night were independently associated with being female compared to male (aOR 2.22; 95%CI 1.55-3.17 and aOR 1.97; 95%CI 1.53-2.54, respectively) and having underlying comorbidities compared to having no comorbidity (aOR 1.98; 1.34-2.93 and aOR 2.06; 95%CI 1.43-2.97, respectively), while forgetfulness (aOR 2.10; 95%CI 1.50-2.95) and palpitations (aOR 2.23; 95%CI 1.44-3.44) were only associated with being female. Additionally, breathlessness at rest (aOR 3.16; 95%CI 1.57-6.38), fits of coughing (aOR 2.93; 95%CI 1.78-4.82), chest tightness (aOR 2.88; 95%CI 1.79-4.63), breathlessness after minimal exertion (aOR 3.05; 95%CI 1.99-4.67), trembling (aOR 4.50; 95%CI 1.97-19.27), coughing when lying down (aOR 2.92; 95%CI 1.67-5.08); confusion (aOR 2.10; 95%CI 1.38-3.18) were associated with having at least one comorbidity.

After considering strength of association, prevalence in cases and control group and effect size, we determined symptoms that were associated with long COVID to be related to three clusters (**Table 3**). Sensory symptoms in order of effect size included loss of taste (aOR 39.69; 95%CI 15.50-101.61), loss of smell (aOR 33.88; 95%CI 14.45-79.45), loss of appetite and blurred vision. Neurological symptoms included forgetfulness, short-term memory loss (aOR 5.66; 95%CI 3.31-9.67) and confusion/brain fog. Cardiorespiratory symptoms included chest tightness/pain, unusual fatigue, breathlessness after minimal exertion (aOR 3.52; 95%CI 2.20-5.62), breathlessness at rest (aOR 3.51; 95%CI 1.68-7.32) and palpitations. The remaining symptoms that were no longer significant in the logistic regression model (at the p≤0.001 level) included trouble trying to form words, coughing when lying down, difficulty swallowing, twitching of fingers and toes, trembling, problems sleeping through the night, fits of coughing and metallic taste.

**Table 3.** Multivariable logistic regression odds ratio (OR) and adjusted odds ratio (aOR) estimates of symptoms of interest.

| Symptom group / symptom | Total<br>N (%)<br>n=1,233 | Cases<br>N (%)<br>n=140 | Control group<br>N (%)<br>n=1,160 | P-value | OR | 95% CI | aOR* | 95% CI | P-value |
| --- | --- | --- | --- | --- | --- | --- | --- | --- | --- |
| **Sensory** |  |  |  |  |  |  |  |  |  |
| Loss of taste | 31 | 24 (17.1%) | 7 (0.6%) | <0.001 | 34.08 | 14.37-80.80 | 39.69 | 15.50-101.61 | <0.001 |
| Loss of smell | 35 | 26 (18.6%) | 9 (0.8%) | <0.001 | 29.17 | 13.34-63.76 | 33.88 | 14.45-79.45 | <0.001 |
| Loss of appetite | 87 | 20 (14.3%) | 67 (5.8%) | <0.001 | 2.72 | 1.59-4.64 | 3.15 | 1.74-5.68 | <0.001 |
| Blurred vision | 99 | 19 (13.6%) | 80 (6.9%) | 0.005 | 2.12 | 1.24-3.62 | 2.61 | 1.47-4.66 | 0.001 |
| Metallic taste in the mouth | 64 | 14 (10.0%) | 50 (4.3%) | 0.003 | 2.47 | 1.33-4.59 | 2.18 | 1.12-4.26 | 0.022 |
| **Neurological** |  |  |  |  |  |  |  |  |  |
| Short-term memory loss | 94 | 29 (20.7%) | 65 (5.6%) | <0.001 | 4.40 | 2.73-7.11 | 5.66 | 3.31-9.67 | <0.001 |
| Confusion/brain fog/trouble focusing attention | 209 | 39 (27.9%) | 170 (14.7%) | <0.001 | 2.25 | 1.50-3.37 | 2.82 | 1.81-4.38 | <0.001 |
| Forgetfulness | 269 | 49 (35.0%) | 220 (19.0%) | <0.001 | 2.30 | 1.58-3.35 | 2.62 | 1.74-3.95 | <0.001 |
| Trembling | 28 | 8 (5.7%) | 20 (1.7%) | 0.002 | 3.45 | 1.49-8.00 | 4.27 | 1.72-10.59 | 0.002 |
| Trouble trying to form words | 129 | 22 (15.7%) | 107 (9.2%) | 0.015 | 1.83 | 1.12-3.02 | 2.24 | 1.32-3.81 | 0.003 |
| Difficulty swallowing | 37 | 9 (6.4%) | 28 (2.4%) | 0.007 | 2.78 | 1.28-6.01 | 3.17 | 1.38-7.26 | 0.006 |
| Problems with sleeping through the night | 682 | 85 (60.7%) | 597 (51.5%) | 0.038 | 1.46 | 1.02-2.09 | 1.63 | 1.11-2.39 | 0.013 |
| Coughing when lying down | 80 | 14 (10.0%) | 66 (5.7%) | 0.045 | 1.84 | 1.01-3.37 | 2.27 | 1.18-4.37 | 0.014 |
| Twitching of fingers and toes | 36 | 8 (5.7%) | 28 (2.4%) | 0.025 | 2.45 | 1.09-5.49 | 2.43 | 1.01-5.80 | 0.046 |
| **Respiratory** |  |  |  |  |  |  |  |  |  |
| Breathlessness after minimal exertion | 154 | 36 (25.7%) | 118 (10.2%) | <0.001 | 3.06 | 2.00-4.67 | 3.52 | 2.20-5.62 | <0.001 |
| Breathlessness at rest | 46 | 13 (9.3%) | 33 (2.8%) | <0.001 | 3.50 | 1.79-6.81 | 3.51 | 1.68-7.32 | 0.001 |
| Chest tightness/pain | 121 | 26 (18.6%) | 95 (8.2%) | <0.001 | 2.56 | 1.59-4.11 | 2.94 | 1.75-4.93 | <0.001 |
| Unusual fatigue/tiredness after exertion | 258 | 55 (39.3%) | 203 (17.5%) | <0.001 | 3.01 | 2.10-4.42 | 2.72 | 1.82-4.07 | <0.001 |
| Fits of coughing | 94 | 19 (13.6%) | 75 (6.5%) | 0.002 | 2.27 | 1.33-3.89 | 2.12 | 1.19-3.75 | 0.010 |
| **Cardiovascular** |  |  |  |  |  |  |  |  |  |
| Palpitations | 163 | 30 (21.4%) | 133 (11.5%) | 0.001 | 2.11 | 1.35-3.28 | 1.93 | 1.20-3.10 | <0.001 |

\*Adjusted for age group, sex, ethnicity, workplace setting and comorbidity status
~Missing observations: 23

The adjusted odds ratios of the three identified clusters of symptoms within the sensory, neurological and cardiorespiratory systems are presented in **Table 4**. Two-thirds (67%) of cases had at least one or more symptoms of the three clusters (versus 44% controls) while one-fifth (20%) reported to having at least one symptom from all three clusters (versus 5% controls) (**Figure 2**). The sensory cluster had the highest association with being a case (aOR 5.25, 95% CI 3.45-8.01).

**Table 4.** Adjusted odds ratios (aOR) estimates using multivariable logistic models of symptoms of interest grouped by cluster.

| Cluster | Total<br>N (%)<br>n=1,233 | Cases<br>N (%)<br>n=140 | Uninfected<br>N (%)<br>n=1,160 | OR | 95% CI | P-value | aOR | 95% CI | P-value |
| --- | --- | --- | --- | --- | --- | --- | --- | --- | --- |
| **Sensory**<br>*Loss of smell OR loss of taste OR*<br>*blurred vision OR loss of appetite* | 191 | 53 (37.9%) | 138 (11.9%) | 4.51 | 3.07-6.63 | <0.001 | 5.25 | 3.45-8.01 | <0.001 |
| **Neurological**<br>*Forgetfulness OR confusion OR short-*<br>*term memory loss* | 323 | 59 (42.1%) | 264 (22.8%) | 2.47 | 1.72-3.55 | <0.001 | 2.77 | 1.87-4.11 | <0.001 |
| **Cardiorespiratory**<br>*Unusual fatigue OR palpitations OR*<br>*breathlessness after minimal exertion*<br>*OR chest tightness/pain OR breathless*<br>*at rest* | 446 | 72 (51.4%) | 374 (32.4%) | 2.23 | 1.56-3.17 | <0.001 | 2.06 | 1.42-3.01 | <0.001 |

\* Adjusted for age group, sex, ethnicity, workplace setting and comorbidity status
~Missing observations: 23

**Figure 2.**
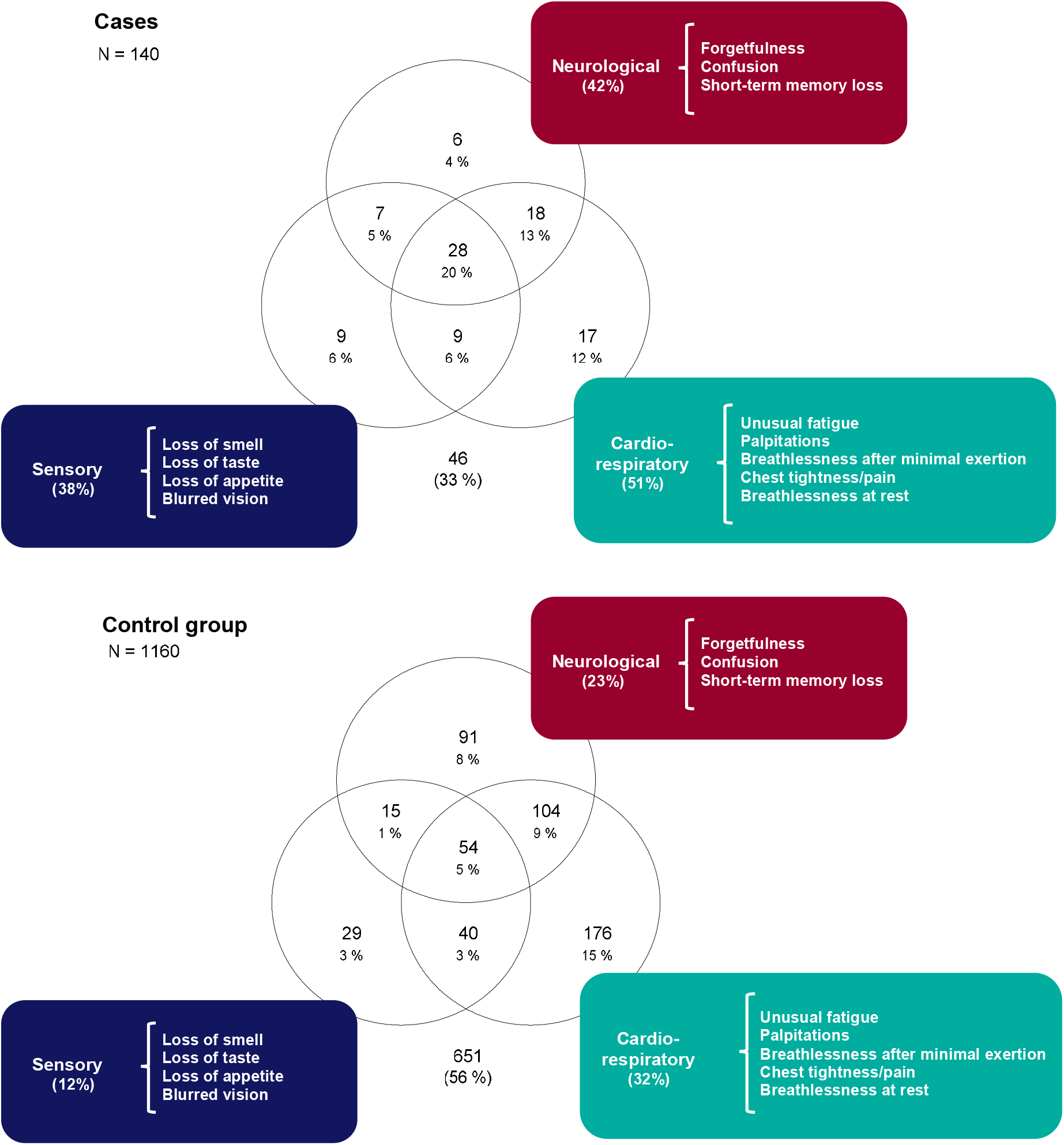
Venn diagrams representing the most common systems affected among cases compared to the control group.

## Discussion

### Summary of findings

We followed-up a unique prospective cohort of clinical and non-clinical healthcare professionals who were regular assessed for COVID-19 since the start of the pandemic. We identified 140 symptomatic cases with mild-to-moderate COVID-19, of whom 3% reported persistent symptoms and a further 11% reported episodic symptoms at a median of 7.5 months after infection. In the month prior to questionnaire completion, cases reported a higher frequency of symptoms affecting the senses, neurological and cardiorespiratory systems than 1,160 asymptomatic, seronegative controls. While cardiorespiratory symptoms including unusual fatigue, breathlessness at rest/after minimal exertion, chest tightness/pain and palpitations were most commonly reported by around half of cases, sensory symptoms had the strongest association, reported by 5 times as many cases as controls. Anosmia (19%) and ageusia (17%) in particular were each >30 times more likely to be reported among cases than controls. Importantly, we found high rates of mental health symptoms which were equally common among cases and controls, highlighting the global negative impact of the current pandemic on then mental health on healthcare workers.

### Comparison with literature

There is currently no accepted case definition for symptoms or duration of long COVID.^7, 17^ Consequently, it is difficult to estimate the prevalence or characteristics of this new condition. Most of the early data on persistent symptoms emerged from follow-up of hospitalised adults with COVID-19, who had more severe illness and, consequently, reported a higher prevalence of ongoing symptoms. One Italian study of 143 patients, for example, who were predominantly male (63%) and with a median age of 57, found 87% of patients with COVID- 19 had at least one residual symptom 36 days after hospital discharge,^15^ while a Canadian study of 78 patients, with similar sex ratio, mean age of 62 and comorbidity prevalence of 41%, reported >75% of patients experienced adverse outcomes three months after onset of symptoms.^18^ In both studies, worsened quality of life and dyspnoea were the most frequently reported adverse outcomes, reported by 40-50% of patients. Fatigue was also common (50%), followed by joint paint (27%) and cough (23%) Notably, persistent symptoms were similar in those with and without pre-existing comorbidities.^18^ Preliminary findings from a large cohort of hospitalised patients found ongoing symptoms reported by 93% of patients, with dyspnea (77%) and fatigue (56%) being most commonly reported at a median of 7 months after illness. Another UK study also found a very high proportion of patients (84%) with at least one residual symptom at follow-up after hospital discharge; this study found that females were significantly more likely to report residual symptoms than males, including anxiety, fatigue and myalgia.^19^ Taken together, these studies of hospitalised patients with COVID-19 are likely to identify high rates of persistent symptoms because of the older age, higher comorbidity prevalence and more severe disease of cases.^20^

Data on sequelae in adults with mild-to-moderate COVID-19 are more limited. In the UK, the app-based COVID Symptom Study known as ZOE found that 13.3% of COVID-19 cases had persistent symptoms lasting >28 days, reducing to 4.5% after 8 weeks and 2.3% after 12 weeks.^14^ Increasing age, female sex, hospital attendance and asthma were independently associated with symptoms lasting beyond 28 days.^14^ By comparison, one-fifth of SARS-CoV- 2 positive participants in the COVID Infection Survey (CIS) conducted by the UK Office for National Statistics (ONS) reported symptoms lasting for longer than 5 weeks and one-tenth exhibiting symptoms for more than 12 weeks.^21^

A major limitation with all such studies, as highlighted by our study, is the lack of a control group to allow attribution of specific symptoms to long COVID. This is exemplified by reported long COVID symptoms such as headache (44%) and hair loss (25%) which were among the top five prevailing symptoms in a systematic review of long COVID.^5^ In our cohort, hair loss was much rarer and not significantly different among cases and controls (9.3% vs 7.4%, respectively), while headache was comparably common (46% cases vs.47% controls).

One US study surveyed 357 COVID-19 cases (9 of whom were hospitalised, 2.5%), 5,497 SARS-CoV-2 negative controls, and 19,095 non-tested individuals and found that 42.3% of cases had at least one persisting symptom at 30 days compared to 13.3% and 8.6%, respectively, with 14.8% still have at least one symptom after 90 days.^9^ This survey found a higher risk of persistent symptoms in those who were initially more ill. The most significant persisting symptoms in those with COVID-19 were anosmia, ageusia, difficulty concentrating, dyspnoea, memory loss, confusion, chest pain, and pain with deep breaths.^9^

The findings are similar to our cohort. Interestingly, while 14% of those with COVID-19 in our cohort reported on-going symptoms after 6 months, most of their symptoms were non-specific and those with more typical persisting symptoms associated COVID-19 such as anosmia or ageusia did not consider them persistent symptoms associated with their initial COVID-19. It was only through specific questioning of individual symptoms that they reported the true range of their symptoms after COVID-19, highlighting the limitations of surveys that rely on self-reporting of symptoms.

The occurrence of anosmia and ageusia after viral infections is not new and occurs through inflammatory reaction of the nasal musosa.^22^ With COVID-19, these features are now included in the case definition for acute COVID-19 and are most common among females. While most individuals recover within a few weeks, there are increasing reports of such symptoms persisting for many months after the acute illness,^23^ as reported in our and the US cohort of individuals with mild-to-moderate COVID-19.^9^ Further follow-up studies are needed to assess the pathogenesis and long-term outcomes of sensory alterations associated with COVID-19.

As with the US survey, we also found a significantly higher prevalence of cardiorespiratory symptoms among cases than controls, although unusual fatigue/tiredness after minimal exertion, breathlessness after minimal exertion and palpitations were also relatively frequently reported by controls (>10%). Not surprisingly, given the predilection of SARS-CoV-2 for the respiratory tract, hospitalised cases (a proxy for severe illness) and mild-to-moderate cases with predominantly respiratory symptoms are reported to be more likely to develop persistent respiratory symptoms at follow-up.^9^

The mechanisms underlying long COVID are largely unknown. It is likely that on-going inflammation, which may persist for several weeks after acute infection, contributes to persistent symptoms affecting specific organs such as the respiratory or cardiovascular systems.^24^ For neurological complications, post-viral, immune-mediated disruption of the autonomic nervous system has been implicated.^25^ It has been postulated that autoreactive T cells and SARS-CoV-2 antibodies that persist after viral clearance may cross-react with host self-antigens including those found in the central nervous system which could manifest as neurological symptoms.^26^ Moreover, there is a precedent for long-term neuropsychiatric consequences after viral infection including post-viral fatigue which is well-described in the literature, including in previous pandemics such as the 1918 Spanish influenza where some patients were diagnosed with encephalitis lethargica post-infection, and, more recently, chronic myalgia, fatigue and disordered sleep following the 2003 SARS epidemic.^27, 28^

Reassuringly, although we identified high rates of mental health symptoms among cases, the inclusion of a well-defined control group found almost identical rates among asymptomatic, seronegative individuals. These findings demonstrate the high toll of the pandemic on clinical and non-clinical healthcare workers that is not necessarily attributable to the infection itself. Some features, however, whilst not infrequent among controls, were significantly more common among cases, including confusion/brain fog/trouble focusing attention, short-term memory loss and forgetfulness. That they persist for more than six months after acute infection in our cohort is concerning and requires further study, including intervention to improve long-term outcomes. Similarly, as with other viral infections, some individuals with COVID-19 will develop chronic fatigue syndrome,^8^ and it is important that these individuals are identified quickly to allow early interventions to improve long-term outcomes.

With respect to the other symptoms, the use of a case-control methodology provides some reassurance that gastrointestinal, rheumatological, dermatological, gynaecological and other non-specific symptoms reported in the literature are equally prevalent among cases and controls.

### Strengths and limitations

The strength in this study lies in the early establishment of a prospective cohort of healthy adults with monthly follow-up including blood sampling and questionnaire completion as well as a high response rate of 78%. Our findings highlight the importance of including a representative cohort of cases to assess long-term outcomes of COVID-19 as well as appropriate controls to estimate the relative prevalence of self-reported symptoms to accurately define this new syndrome. There are, however, some limitations. The participants were clinical and non-clinical healthcare workers who may not be representative of the general population. Additionally, compared to controls, those with confirmed COVID-19 were more likely to be patient-facing, frontline clinical healthcare workers, who have also been working throughout the pandemic and, therefore, more likely to suffer from adverse physical, mental and emotional outcomes, including post-traumatic stress and burnout.^29^ Another potential bias may result from participants already being aware of their SARS-CoV-2 RT-PCR and antibody status when completing the questionnaire. There may be also be some recall bias in that cases may have spent more time thinking about their symptoms than the asymptomatic control group. All these limitations, however, would increase the prevalence of symptoms – especially mental health symptoms – among cases compared to controls. The lack of a statistically significant difference for many of the symptoms, therefore, strongly suggest that these features are not associated with long COVID.

## Conclusions

SARS-CoV-2 infection is associated with a broad spectrum of clinical disease, ranging from mild, transient respiratory illness to multi-organ failure requiring intensive care support.

Recovery from infection will, therefore, vary according to severity of the initial illness and the organs affected. Assessment of an established cohort of healthy adults with mild-to-moderate COVID-19 and a well-defined group of asymptomatic, antibody negative controls identified a small group of patients with very specific persistent symptoms more than six months after infection. Our study adds to the evidence-base for the long-term effects of COVID-19 in adults with mild-to-moderate COVID-19 who contribute to the vast majority of 120+ million infections worldwide. This information is not only important for clinicians, patients and the public, but also for policy makers and healthcare providers who are investing heavily in long-term provisions for COVID-19 survivors. On-going surveillance will be important to monitor the physical and mental health of our cohort.

## Data Availability

Data not available due to ethical restrictions

## Ethics approval

This study was approved by PHE Research & Public Health Practice Ethics and Governance Group (R&D REGG Ref NR 0190).

## Funding

This study was internally funded by Public Health England with additional support provided by the NIHR Manchester Clinical Research Facility. The study was also carried out at the NIHR Manchester Clinical Research Facility. AH and BP are supported by the NIHR Manchester Biomedical Research Centre. The views expressed are those of the authors and not necessarily those of the NGS, the NIHR, of the Department of Health.

## Patient and Public Involvement

Patients or the public were not involved in the design, or conduct, or reporting, or dissemination plans of our research

## Notes

### Competing Interest Statement

The authors have declared no competing interest.

